# Machine Learning in the analysis of lethality and evolution of infection by the SARS-CoV-2 virus (COVID-19) in workers of the Mexico City Metro

**DOI:** 10.1101/2021.10.27.21265573

**Authors:** Eréndira Itzel García Islas, Guillermo de Anda Jáuregui, Joaquín Salas Rodríguez, Florencia Serranía Soto

## Abstract

In terms of the number of fatalities, Mexico has been one of the countries most affected worldwide by the pandemic. Using different Machine Learning techniques, some of the first cases of the infection registered in Mexico City (CDMX), the geographical and political center of the country, are analyzed in order to determine the causes of lethality and evolution of infection by the SARS-CoV-2 virus, from April 1 to September 27, 2020 in workers of the Capital Metro.

## Introduction

The Mexico City Metro is a massive public transport system, a metropolitan train type, which serves large areas of the Valley of Mexico and the conurbated area. It has 12 lines with a total length of 226.49 km distributed in 195 stations, 3,333 wagons with a standard capacity of 1,530 people per train; 15,239 direct employees; 1,685 annual trips per passenger and 114.03 million kilometers traveled per wagon per year^1^.

In order to reach a better understanding of the origin, dispersion and evolution of the SARS-CoV-2 virus (COVID-19), which causes Severe Acute Respiratory Syndrome, some of the first cases of infection registered in the country are analyzed. In particular, the one that occurred on february 6, 2020 among the workers of the Mexico City Metro (CDMX).

### Distribution of cases by age and sex

Based on databases analysis with information from the health system of the health care clinics of the Capital Metro, the confirmed cases with the SARS-CoV-2 virus during the period from April 1 to September 27, 2020 are analyzed.

In the aforementioned interval, a total of 4.15 consultations were observed, which correspond to the care of 511 workers diagnosed with the SARS-CoV-2 virus, so an average of 8 consultations linked to symptoms due to the described pathology or associated comorbidities are recorded. In relation to patients infected with the SARS-CoV-2 virus, it is observed that of the 511 confirmed cases with the disease, 152 correspond to women between 21-70 years of age (μ = 44.6,σ = 9.3), 359 to men between 19-73 years of age, while the vulnerable population at risk due to advanced age (60 years and over) included only 19 individuals, that is, 3.7% of the workers analyzed.

Of the patients diagnosed with COVID-19, 436 workers (85.32%) received outpatient treatment, while 75 workers (14.68%) required hospitalization, including 53 men, representing 14.76% of the male population and 22 women, 14.47% of the infected female population, with a percentage of deaths of 1.34% for both sexes and an average of 17 days of hospital stay for women and 28 days for the male sex from the patient’s admission to the unit health. Patients who required hospitalization were referred for care to two clinics, the first concentrating 70.66% and the second 29.34% of cases, respectively.

### Distribution of cases by administrative unit

According to the number of infected by department, administrative unit and level of command, during the aforementioned period, 142 infected workers worked in the Transportation Directorate, 124 in the Fixed Facilities Management, 97 in the Rolling Stock Maintenance Directorate, 60 belonged to the Box Office Coordination,, 18 to the Directorate of Material Resources and General Services, 21 workers held managerial positions without assigned level of command, while 49 held other positions.

### Diagnosis and evolution of the disease

Among the main causes that demanded medical attention in patients diagnosed with the SARS-CoV-2 virus, 627 consultations and/or requests for medications corresponded to the care of infections of the lower and/or upper respiratory tract (acute or chronic bronchitis, pneumonia, tonsillitis, pharyngitis, laryngitis); 495 to symptomatic treatment of fever and/or pain; 477 to the care of gastrointestinal disorders (meteorism, nausea, vomiting, constipation, diarrhea, heartburn, as well as gastritis, colitis and / or duodenitis associated or not with the consumption of drugs); 281 clinical reviews were referred to the treatment of muscle or joint pain; 228 to type 1 and 2 diabetes care; 163 to the control of hypertension problems; 160 to the treatment of gastrointestinal infections (salmonellosis, typhoid fever); 140 care for Chronic Obstructive Pulmonary Disease (COPD); 112 to the care of autoimmune diseases (rheumatoid arthritis, lupus, multiple sclerosis); 106 to the control of hypercholesterolemia and hypertriglyceridemia; 102 to the care of dermatological diseases (psoriasis, impetigo, dermatophytosis, mycosis); 88 to the treatment of neurological diseases; 77 to the care of endocrinological disorders (hypothyroidism, hyperprolactinemia); 69 consultations were intended for the treatment of depression associated or not with the disease; 56 to the treatment of ophthalmological diseases (cataract, glaucoma, superficial lesions of the cornea); 56 to the care of urogenital infections; 21 to the care of cases of heart failure; 20 to the treatment of pulmonary thrombosis; 15 to the attention of anxiety disorders; 46 to the treatment of ventricular arrhythmias and 8 to the care of cases of renal failure; among other conditions with no apparent relationship to the disease (see Table 1).

**Table 1.**
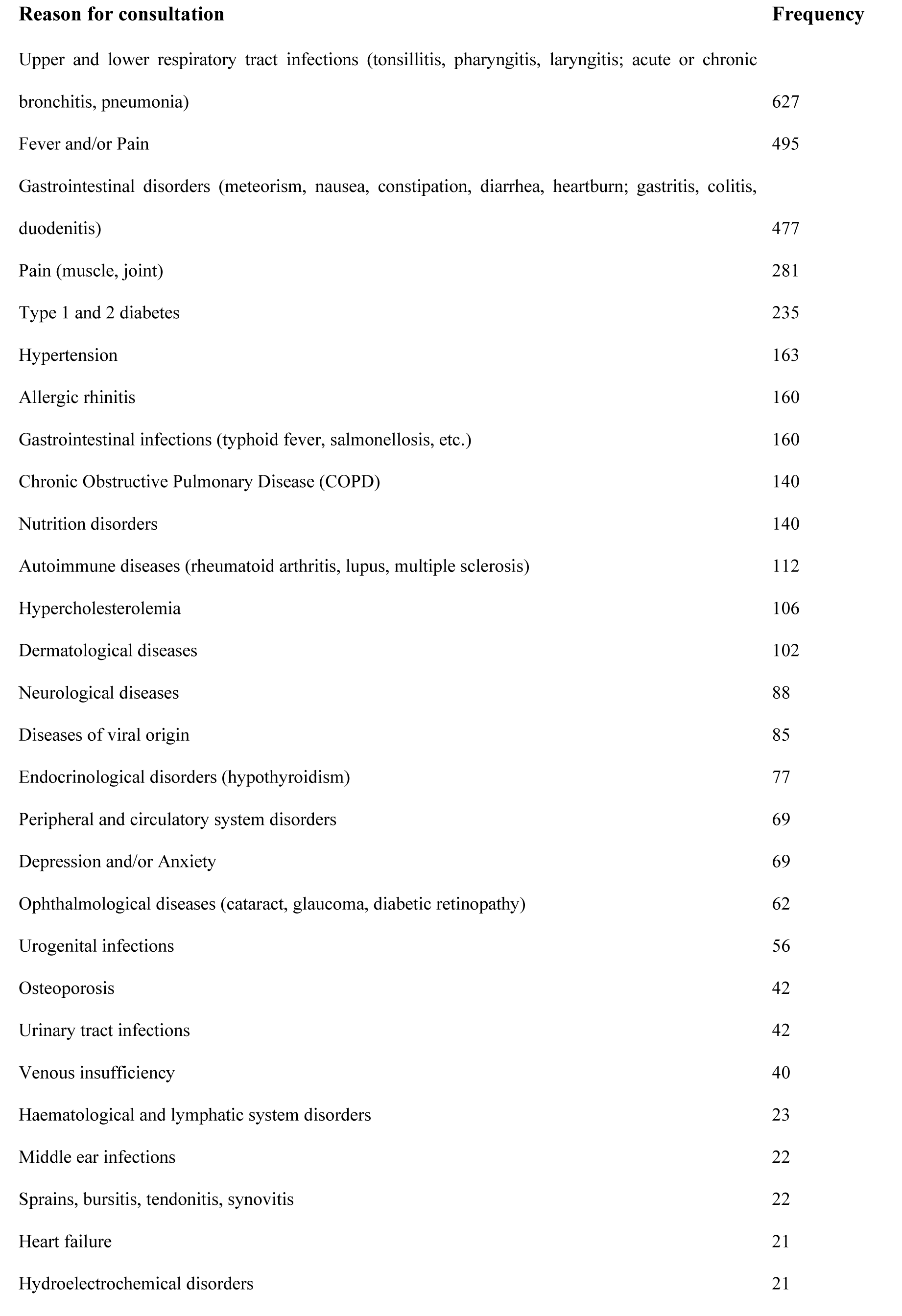
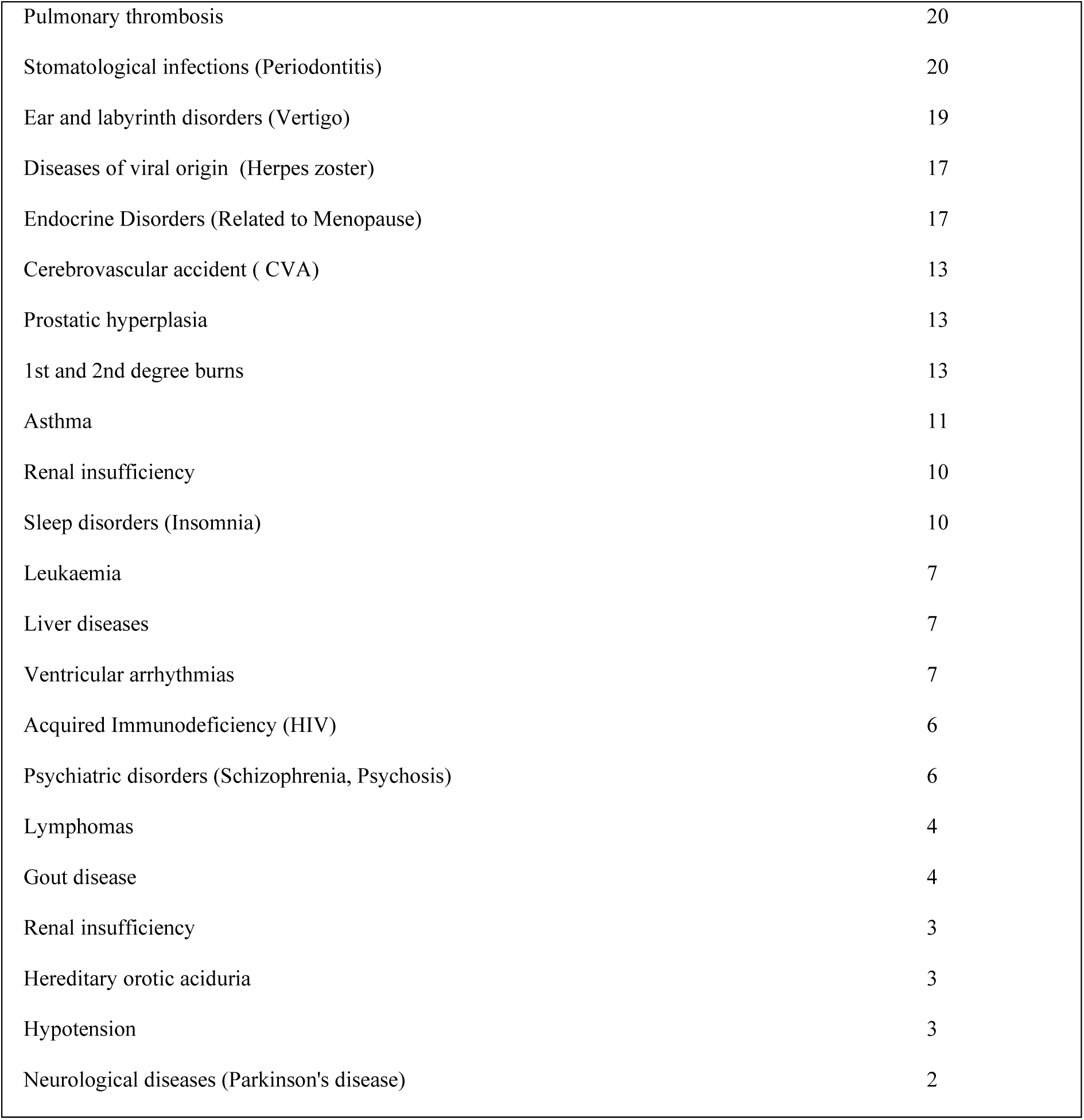
Reason for consultation and frequency of occurrence. Among the reasons for occurrence are the care of respiratory diseases, the presence of fever associated with infectious diseases and the care of gastrointestinal disorders

**Table 2.**
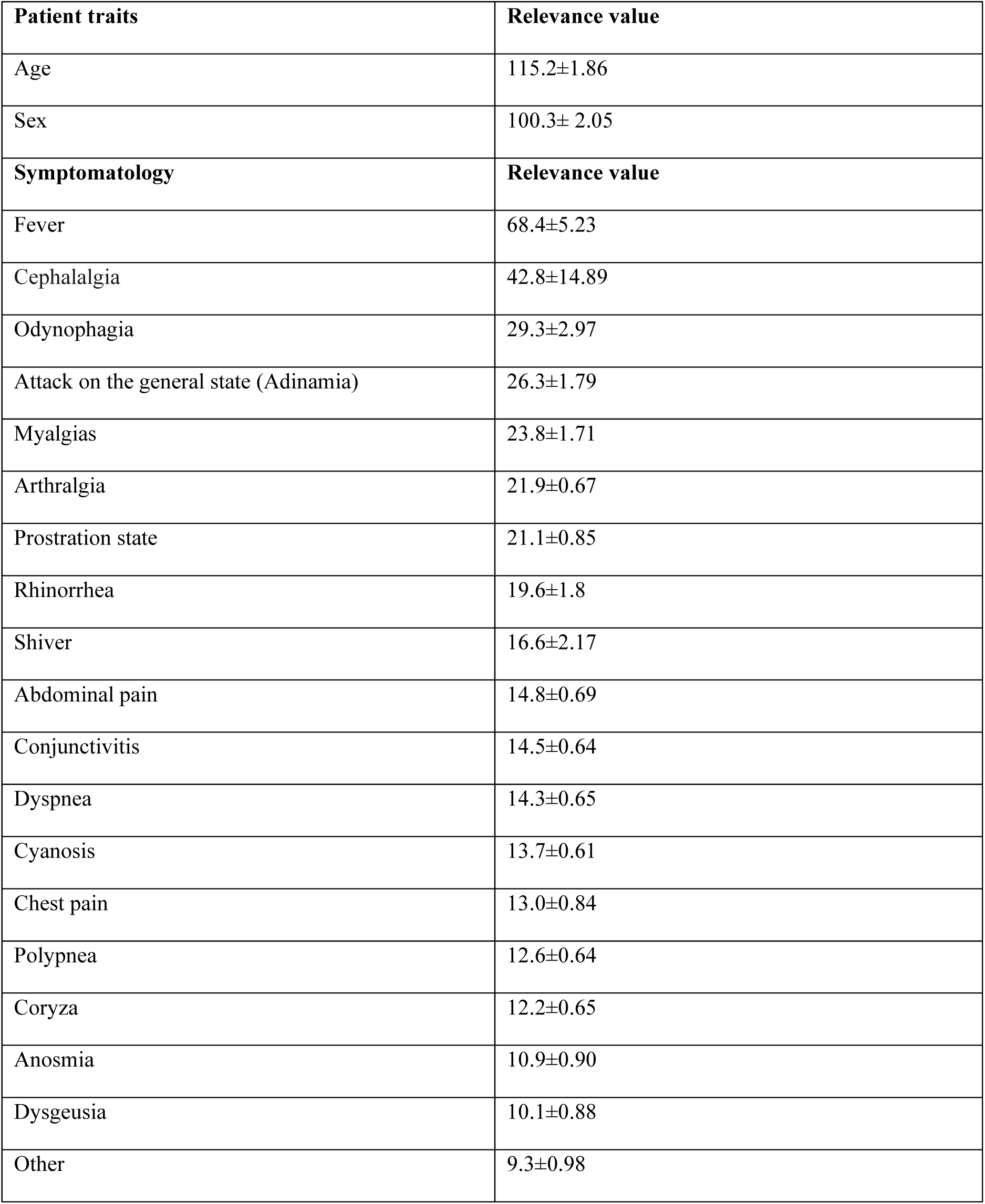
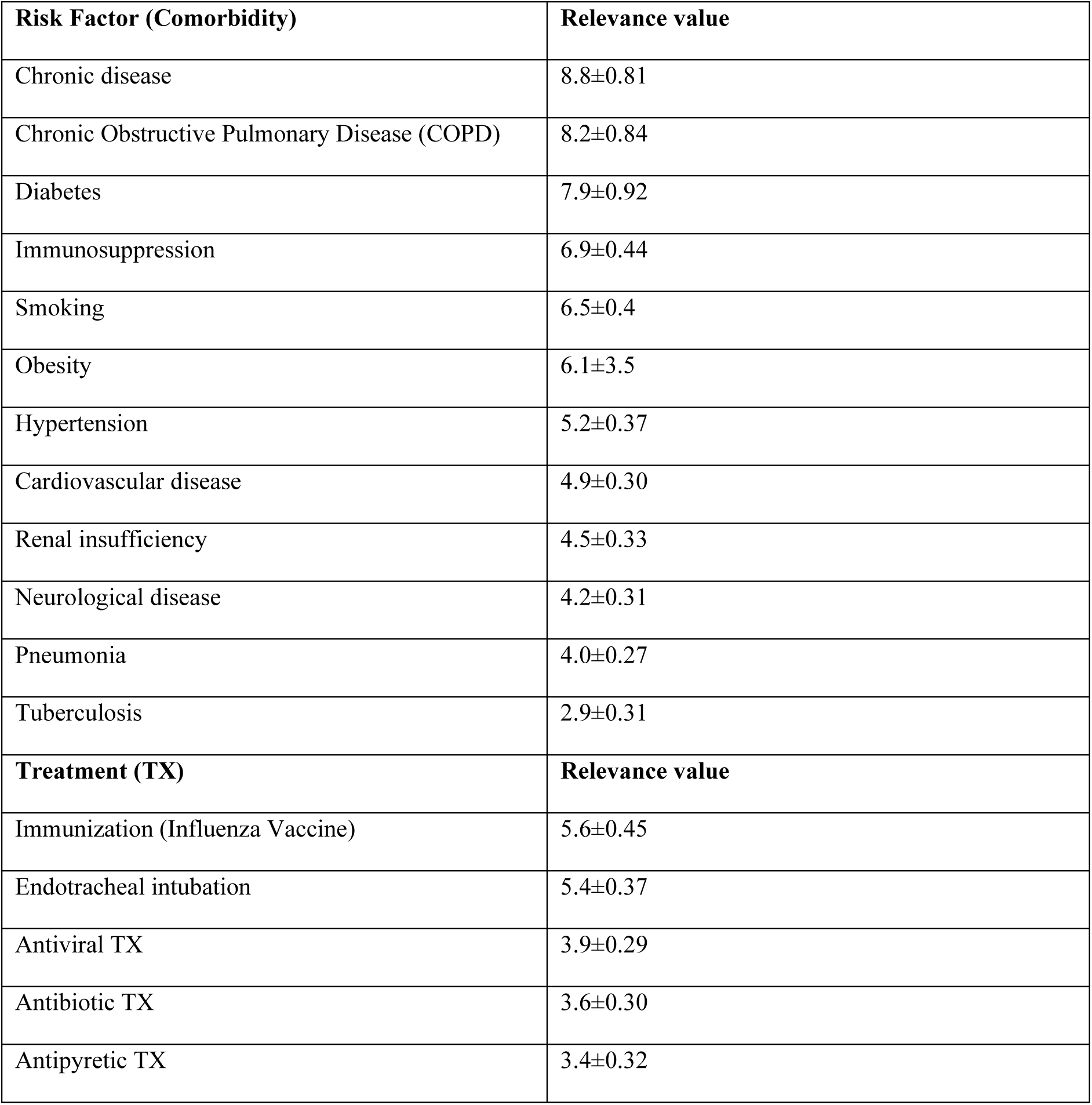
Relevant characteristics determined from neural network training.

### Treatment and evolution

Regarding the type of treatment received, patients were treated with retrovirals such as oseltamivir, amantadine, dolutegravir, lopinavir/ritonavir, abacavir/lamividuine, emtricitabine/tenofovir, antiparasitics such as ivermectin, nitazoxanide, quinfamide and metronidazole while the added infections were treated with azithromycin, levofloxacin, trimethoprim + sulfamethoxazole, cefixime and even immunomodulators such as tocilizumab.

### Comorbidity análisis

Of the total staff infected by the SARS-CoV-2 virus, a total of 238 workers at risk for comorbidity associated with Chronic Noncommunicable Diseases (NCDs) were identified, of which 182 reported a history of respiratory diseases (35.61%), 73 hypertension (14.28%), 66 hypercholesterolemia and / or hypertriglyceridemia (12.91%), 59 Diabetes (11.54%), 55 presented obesity (10.76%), 6 heart diseases (1.17%), 6 kidney diseases (1.17%), 3 liver diseases (0.58%), one worker reported immunosuppression (0.19%) and 6 of them reported tobacco use (1.17%). However, the evolution of the virus in outpatients occurred generally uncomplicated. In the case of hospitalized patients, 11 deaths occurred during the period analyzed.

### Mortality rate by age

When considering cases in age groups of 20 years, it is observed that the mortality rate (case fatality rate) increases progressively as age increases, from 9.70% for the group of 20 to 40 years, 16.50% for the group of 40 to 60 years, and up to 36.8% for the group of 60 to 80 years of age. Figure 1 shows that in the “over 60 years” group, the risk of death is the highest of all age groups. This observation is consistent with the data reported for the general population, both in Mexico and in other countries.

**Figure 1.**
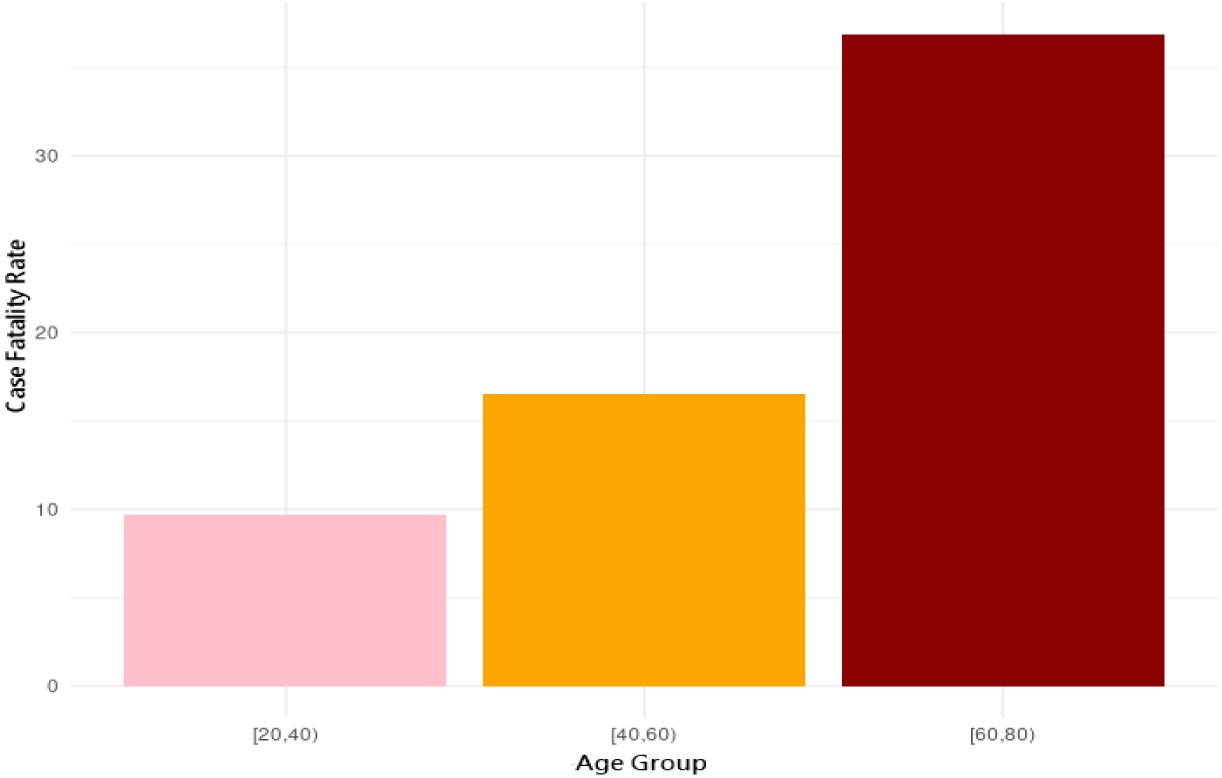
Mortality rate by age group.

### Epidemiological dynamics

From the analysis of the epidemiological dynamics, it is observed that the highest number of confirmed cases during the aforementioned period occurred between epidemiological weeks 17 to 23. On the other hand, there is a peak in the hospitalization curve in week 25 of the year 2020. (Figures 2 and 3)

**Figure 2.**
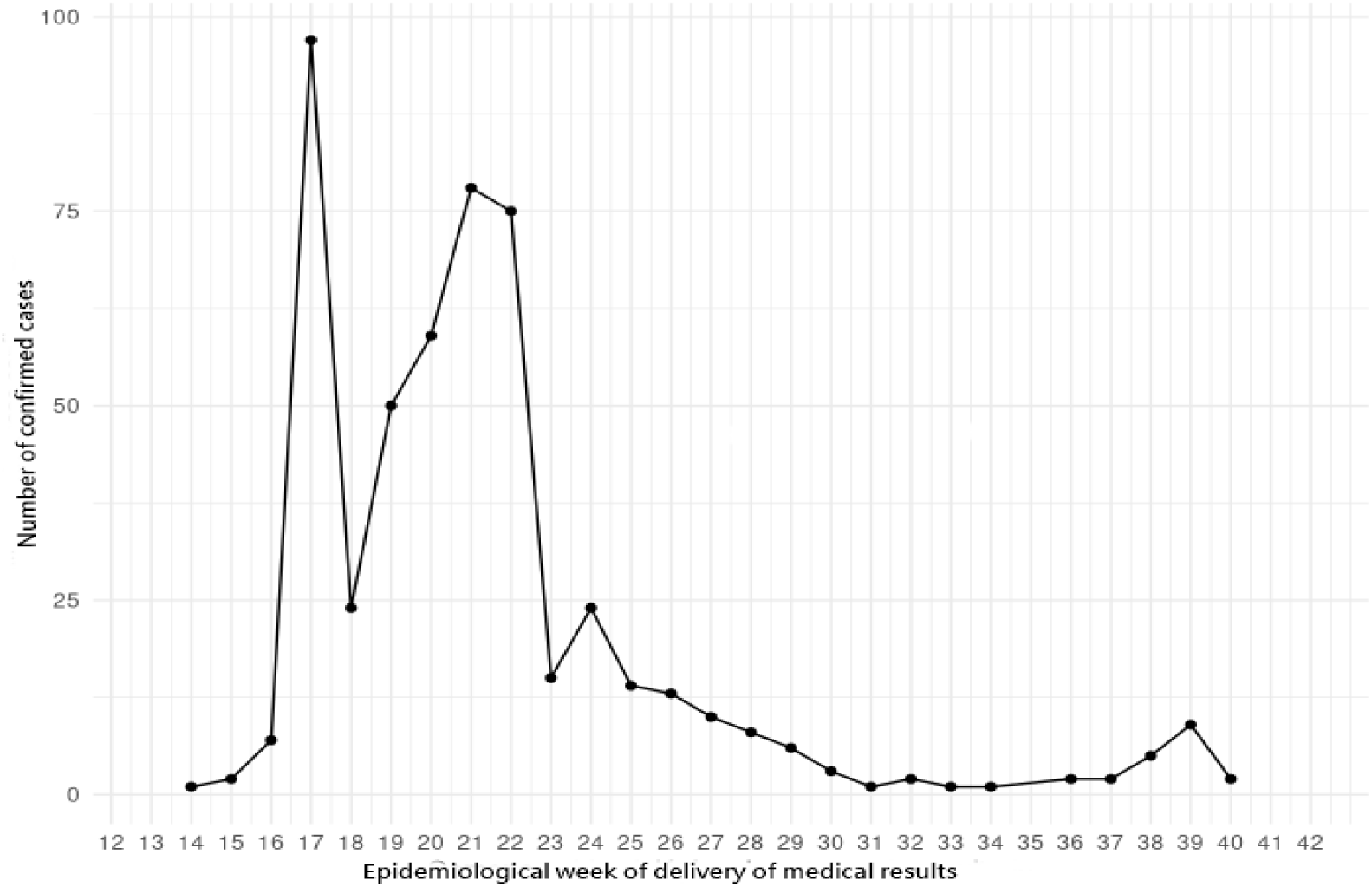
Number of confirmed cases per epidemiological week (Incidence curve considering positive diagnosis date)

**Figure 3.**
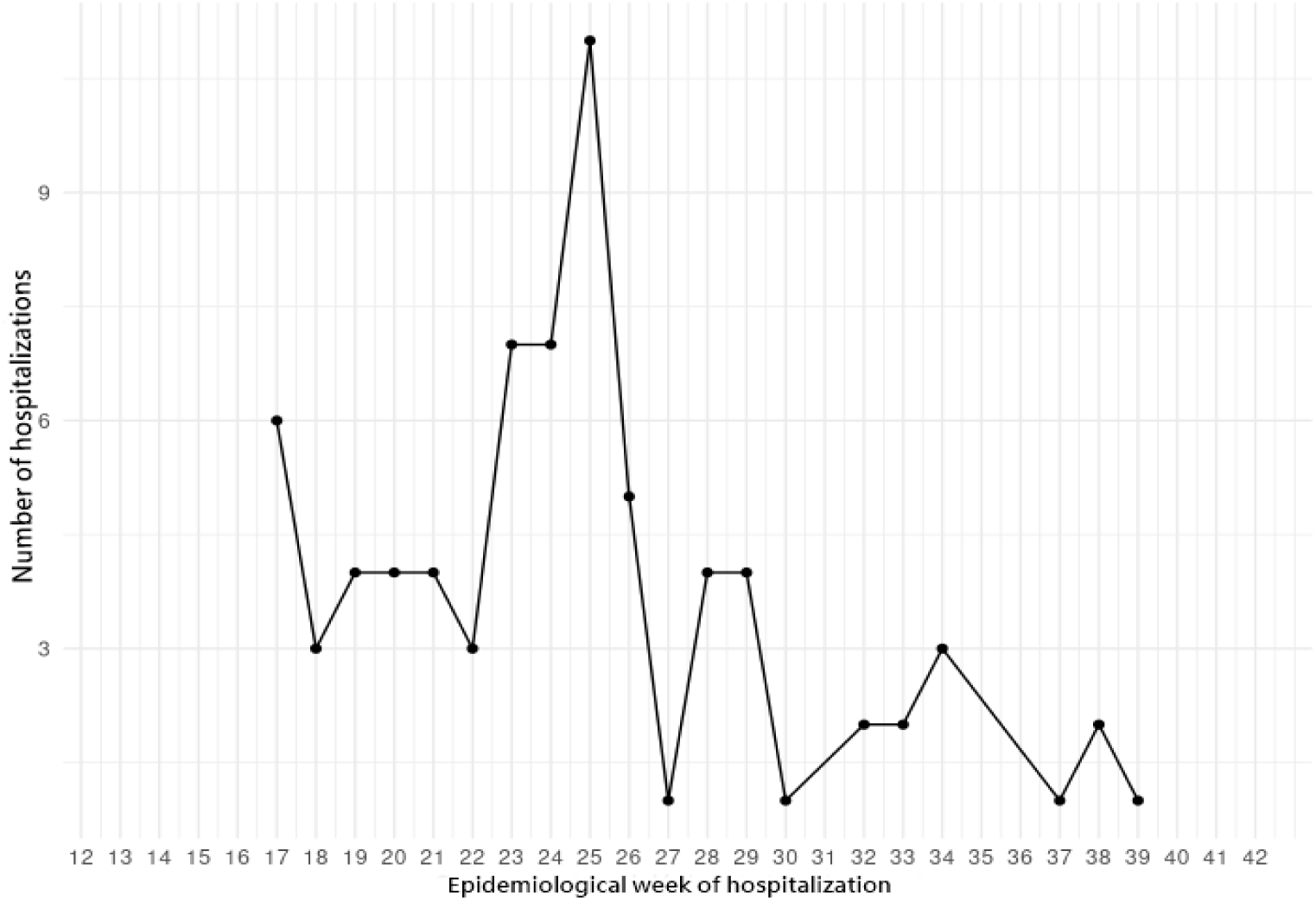
Number of hospitalized patients per epidemiological week (Considering date of hospitalization)

It should be noted that, in general, the curves described coincide with the epidemiological dynamics observed in Mexico City, as well as with that reported by federal authorities from the database of the National Epidemiological Surveillance System (SINAVE).

### Clinical prognosis

In order to determine the lethality and evolution of the disease, the data set described ***C*** = {*c*_1,_ ⋯, *c*_*n*_} was analyzed with 511 positive cases confirmed by COVID-19 in workers of the Metro of the CDMX, as well as the information provided during the face-to-face consultation in the aforementioned health units and the data set for COVID-19 of the National Epidemiological Surveillance System (SINAVE) of the CDMX until December 7, 2020, which contained a total of 145,759 records with 51,137 active patients, of which 32,178 observed improvements in their state of health, while to the aforementioned date 9,785 deaths had been registered. Given that the original dataset presents a total of 209 predictors, it is interesting to distinguish between those patients who recovered from the disease and the deaths that occurred in the aforementioned period.

The cases for which the physician had issued a decision or line of treatment according to the diagnosis and possible evolution of the patient based on the analysis of comorbidity determined, were denoted by *D* = {*d*_1,_ ⋯, *d*_*n*_}. In particular, it is possible to express the different decisions as classes of interest. That is, as *d*_*j*_ ∈ {1, ⋯, *k*} where *k* denotes the number of classes.

Each record contains the results of the clinical intervention derived from the face-to-face interview, laboratory analysis or from previous clinical diagnosis, described in the form of characteristics that are associated with each record as 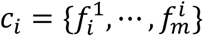.

The objective of this analysis is to determine, independently of the initial characteristics *m*, which are the values of *m*′ with greater relevance that allow to discriminate between the results of interest (recoveries-deaths). To this end, classifiers were built to distinguish between classes of interest.

### Feature Selection

The feature selection process consists of choosing the subset of *m*′ traits of greatest relevance within the existing *m’*s in a dataset, with the aim of classifying an observation into one of the *k* classes, improving prediction performance, as well as providing a better understanding of the underlying process that gave rise to the data.

Among the methodologies and techniques that make the operation of the models described and the selection of traits more efficient, are filter-based methods and enveloping selection methods. The latter use a classifier to evaluate different subsets of traits based on a metric that allows choosing the best representative and use the performance of that characteristic as an evaluation criterion. In this way the most suitable trait for the algorithm is obtained. Filter-based methods, on the other hand, look for statistical relationships between the traits and the target variable and use this score as the basis for selection.

### Experimental configuration

For the case of interest, the number of samples of the original class was equalized and an enveloping selection method called Boruta was used, an algorithm that provides criteria for the selection of relevant attributes through the use of classifiers. Boruta yields a measure of relevance for each of the traits and operates by extending the characteristic base with artificial variables, one for each of the original variables. The algorithm then combines the values of the artificial predictors before running the classifier for the extended system. From a statistical test, Boruta compares the value of the most relevant artificial variable with the original ones. All original variables that result in values of importance greater than those of artificial variables are considered relevant. Due to the random nature of the variables that are combined, additional iterations of the above process can result in different characteristics. The previous mechanism is extended with an election process. The Boruta schema runs *b* times and those features are selected as important 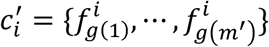 that consistently exceed the decision criteria in each iteration, for a g function that assigns the selected entities to the originals.

### Machine Learning Classification

By employing the *m*′ characteristics considered relevant by the selection process, machine learning classifiers were generated *f*_*h*_ assigning the characteristics 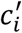 of an example to their corresponding class *d*_*i*_.. In the present analysis, the following classifiers were analyzed:

#### Random Forest

Classifier based on decision trees, in which the training samples are randomly chosen, as well as the predictors used to perform each random division.

#### Neural Networks (Neural Networks NN)

They represent universal mappers, that is, they allow to approximate any function with an arbitrary level of precision. Neural networks present a hierarchical learning model in which each layer projects the inputs in non-linear spaces, possibly of greater dimension. In the last layer, hyperplanes are expected to be able to discriminate between classes.

#### Support Vector Machine (SVM)

In general, Vector Support Machines allow to find an optimal hyperplane that maximizes class separation. In the case of classes that are not linearly separable, SVM uses the projection of the data to a nonlinear and higher dimensional space, through kernel functions, product point, and nonlinear functions that meet Mercer’s criterion as a measure of likelihood.

#### Extreme Gradient Boosting (XGB)

It produces a predictive prototype in the form of a set of decision trees. It constructs the model in a staggered way analogous to other boosting methods and generalizes them allowing the arbitrary optimization of a differentiable loss function.

For the present analysis, the selection of characteristics was executed ten times and those traits that were considered relevant in each iteration were retained, using the technique known as Random Forest.

From the aforementioned classifiers, a system of selection based on neural networks was defined that takes as input the output of each classifier and generates the medical prognosis. Available data were divided into subsets of training, assessment and testing. To evaluate performance, reception performance characteristics (ROC) and precision recovery curves (PR) were used. It is important to note that the results are provided as area under the curve (AUC) either ROC or PR, and uncertainty is limited by iteration of the above procedure.

In this way, the algorithm obtained 39 important characteristics. The traits or factors determined from the selection algorithm suggest that the values with the greatest relevance that allow discriminating between the results of interest (recoveries-deaths) correspond to the following variables:

This allows us to affirm that the mortality rate associated with infection by the SARS-CoV-2 virus (case fatality rate) is affected by different risk factors related to traits, comorbidities and pharmacological treatment received.

Lethality increases progressively as age increases. While the probability of contagion is the same for both sexes, the mortality rate in infected men is almost double the mortality rate in the female population. Thus, if the patient is male, over 60 years of age, the risk of mortality increases with a preponderance of 100.31 and 115.2, respectively, so even when the role of sex in the prevalence of infection by the SARS-CoV-2 virus is unknown, the evolution towards negative pictures of the disease increases significantly in the male population. This allows us to suggest the existence of a factor of hormonal origin that affects the propensity of the male sex to suffer severe effects of the disease, as well as to present a greater risk of lethality.

On the other hand, the most important common symptoms related to infection by the SARS-CoV-2 virus are, in order of prevalence: Fever, cephalalgia, odynophagia, attack on the general state (adinamia), myalgia, arthralgia, prostration, rhinorrhea, chills, abdominal pain, conjunctivitis, dyspnea, cyanosis, chest pain, polypnea, coryza, anosmia, dysgeusia, among others. While the risk factors related to the pre-existence of pathologies that increase lethality correspond to: Chronic Obstructive Pulmonary Disease (COPD), Diabetes, Hypertension, Immunosuppression, Smoking, Obesity, Cardiovascular disease, Kidney failure, Neurological disease, Pneumonia and Tuberculosis. Regarding the treatment received, immunization through the influenza vaccine close to infection with the SARS-CoV-2 virus increases the risk of lethality therefore, it is suggested to evaluate the existence of a possible interference in the immune response to the coronavirus for the development of complications from infection by the SARS-CoV-2 virus in the adult population.

Likewise, patients who require endotracheal intubation have a higher risk of lethality than those receiving antiviral or antibiotic treatment, while the lowest risk is located in cases receiving outpatient treatment to which antipyretic is administered in combination with any of the pharmacological treatments described above.

### Related literatura

Lalmuanawna et.al (Lalmuanawma, Hussain, & Chhakchhuak, 2020) present a compendium of current Machine Learning applications related to SARS-CoV-2 virus (COVID-19) infection. Their review suggests that there is a vast body of evidence for ML research to play a leading role in the evolution and outcomes of the epidemic. In this context, Sharma (Sharma, 2020) trains convolutional neural networks (NNBs) from images from healthy people (618) and infected with influenza (175), pneumonia (224) and COVID-19 viruses to predict the results of the qRT-PCR test of pulmonary computed tomography (CT) scans. Similarly, Mei et al. (Mei, et al., 2020) explore alternatives to the qRT-PCR test by using AI techniques for rapid diagnosis of COVID-19 using data from Computed Tomography (CT), clinical symptoms, exposure history, and patient laboratory tests. They then employ a hybrid technique that makes use of machine learning methods such as Random Forest (RF), Multilayer Perceptron (MLP), and Vector Support Machines (SVM) as classifiers.

In relation to the problem of interest of the present study, the researchers have analyzed cases of patients who have survived the SARS-CoV-2 virus to discriminate between patients in general, those seriously ill and deceased. In this context, Yan et al. (Yan, et al., 2020) present an ML method that allows inferring clinical prognosis from three blood markers. Lactic dehydrogenase (LDH, associated with tissue degradation in pulmonary atrophies), lymphocytes (a subtype of white blood cells that are part of the innate immune system) and highly sensitive C-reactive protein (hs-CRP, which increases blood flow during the inflammatory process and the presence of infections). Additionally, researchers have developed AI techniques for the selection of risk factor characteristics and medical prognosis. Nemati et al. (Nemati, Ansary, & Nemati, 2020) implemented a survival analysis using 1,182 COVID-19 cases in order to predict their recovery time from characteristics associated with age and sex, optimizing results with an XGB classifier. Schwab et al. (Schwab, Schutte, Dietz, & Bauer, 2020) implemented an ML algorithm to predict the outcome of qRT-PCR tests as well as to determine whether a confirmed positive case will require hospitalization or intensive care.

In a large-scale experiment, Pourhomayoun and Shakibi (Pourhomayoun & Shakibi, 2020) assessed mortality risk for COVID-19 patients from the analysis of 117,000 patient records from 76 countries. To this end, they trained different ML classifiers, including SVM, artificial neural networks (ANN), RF, logistic regression (LR) and k-Nearest Neighbors (kNN), from where they observed that ANN presented the best performance.

Souza et al. (Souza, Santos, Silva, & Guidoni, 2020) predicted the COVID-19 test outcome in a closed group of 13,690 patients from a set of AI techniques and ML methods, including LR, Linear Discriminant Analysis (LDA), Naive Bayes (NB), kNN, Decision Trees (DT), XGB, and SVM. From an international research effort that includes the United States and the European Union, (Bertsimas, and others, 2020) a method of mortality risk assessment was developed from XGB-based classifiers, clinical and laboratory predictors. Among the critical risk factors, age, reduced oxygen saturation, high levels of PCRhs, as well as the presence of creatinine, urea nitrogen and blood glucose were found. In the case study, an enveloping feature selector was employed, and unlike the studies cited, the role of regional data was emphasized.

### Discussion and Conclusion

The CDMX Metro as a public transport system, intended for the transfer of 5 million 72 thousand 384 daily travelers in one of the largest cities in the world, offers a study scenario that can be used to understand the origin and expansion of the epidemic nationwide. In particular, it seems to explain why Mexico represents one of the countries with the highest mortality from COVID-19 in the world. The sample population reflects an age group with relevant risk, while its figures of contagion, hospitalization and mortality coincide with those of the rest of the population. In this sense, by coinciding the associated risk factors for death and improvement with those applied to the open population, the prediction values thrown by the algorithm can be transferred without risk of ambiguity to the population of the Metro of the CDMX, therefore, the experimental results obtained through this study suggest that the population of the Metro can be considered as a representative group of the origin, dispersion, and evolution of the COVID-19 pandemic in Mexico City.

According to the classification algorithm, the lethality and evolution of the disease are affected by 39 risk factors related to traits, comorbidities, as well as the treatment received. The lethality increases progressively as age increases, so it is suggested to evaluate the role of sex in the prevalence of infection by the SARS-CoV-2 virus, because the evolution towards negative pictures of the disease occurs mostly in the male population, which suggests the possible existence of a factor of hormonal origin.

Approximately 18 common symptoms related to SARS-CoV-2 virus infection were identified, in order of prevalence: Fever, headache, odynophagia, attack on the general condition (adinamia), myalgia, arthralgia, prostration, rhinorrhea, chills, abdominal pain, conjunctivitis, dyspnea, cyanosis, chest pain, polypnea, coryza, anosmia, dysgeusia, among others. The risk factors found by the algorithm, which increase lethality, are related to the pre-existence of pathologies such as: Chronic Obstructive Pulmonary Disease (COPD), Diabetes, Hypertension, Immunosuppression, Smoking, Obesity, Cardiovascular Disease, Kidney Failure, Neurological Disease, Pneumonia and Tuberculosis. Regarding the treatment received, immunization through the influenza vaccine close to infection by the SARS-CoV-2 virus, increases the risk of lethality, so it is suggested to evaluate the existence of a possible interference in the immune response to the coronavirus for the development of complications from infection by the SARS-CoV-2 virus in the adult population. Likewise, patients who require endotracheal intubation present a higher risk of lethality than those receiving antiviral or antibiotic treatment, while the lowest risk is located in cases receiving outpatient treatment to which antipyretic is administered in combination with any of the pharmacological treatments described above.

### Future work

As future work, the analysis of the correlation between characteristics of the population in general and their representativeness in the capital’s Metro will continue, in order to support the definition of public policies aimed at reducing the adverse effects of the pandemic in Mexico City.

## Data Availability

All the data produced in this work are contained in the manuscript

https://www.sinave.gob.mx/

https://www.gob.mx/salud/

## Footnotes

1 Collective Transportation System, Mexico City, Mexico: https://metro.cdmx.gob.mx/operacion/cifras-de-operacion.

## Notes

### Competing Interest Statement

The authors have declared no competing interest.

### Funding Statement

This study did not receive any funding

### Author Declarations

Dataset for COVID-19 from the National Epidemiological Surveillance System (SINAVE) www.sinave.gob.mx

